# Brief Report: Sensory processing phenotypes in Phelan-McDermid Syndrome and *SYNGAP1*-related Intellectual Disability

**DOI:** 10.1101/2020.11.30.20241315

**Authors:** Ariel M Lyons-Warren, Maria C McCormack, J. Lloyd Holder

## Abstract

Sensory processing differences are an established feature of both syndromic and non-syndromic Autism Spectrum Disorders (ASD). Significant work has been done to characterize and classify specific sensory profiles in non-syndromic Autism. However, it is not known if syndromic Autism disorders such as Phelan-McDermid Syndrome (PMD) or *SYNGAP1*-related Intellectual Disability (*SYNGAP1*-ID) have unique sensory phenotypes. Understanding the sensory features of these disorders is important for providing appropriate care and for understanding the underlying mechanisms of the disorders. In this manuscript we use the Short Sensory Profile-2 to characterize sensory features in 41 patients with PMD and 24 patients with *SYNGAP1*-ID and compare their responses to both expected results for typically developing children and published sensory profiles for non-syndromic ASD.

---

Sensory processing differences are a common feature of many neurodevelopmental disorders (Dunn et al. 2016; Smith et al. 2020). For example, hyper and hypo sensitivity are common in non-syndromic Autism Spectrum Disorder (ASD) (Robertson and Baron-Cohen 2017) and complex challenges with sensory processing have been documented in tic disorders (Soler et al. 2019), Fragile X syndrome (Rais et al. 2018), attention deficit hyperactivity disorder (Little et al. 2018) and a limited number of syndromic ASDs (Heald et al. 2019). Not surprisingly, the specific sensory phenotypes and distributions of sensory differences vary across diseases. Thus, characterizing the sensory profiles for specific disorders is important for diagnosis, management and understanding underlying disease mechanisms.

Rare monogenic neurodevelopmental disorders provide a framework for studying common phenotypic features such as sensory processing differences (Soorya et al. 2018). Phelan-McDermid syndrome (PMD) and *SYNGAP1*-related intellectual disability (*SYNGAP1*-ID) are two such examples of rare neurodevelopmental disorders. PMD is caused by disruption of *SHANK3*, usually due to deletion of chromosome 22q13.3 or sometimes *de novo* loss-of-function single nucleotide variants (SNVs) in *SHANK3*. Patients present with neonatal hypotonia, developmental delay often ultimately diagnosed with intellectual disability and often ASD as well as characteristic facial and hand features (Phelan et al. 2018). Similarly, *SYNGAP1*-ID is caused by de novo loss-of-function SNVs in *SYNGAP1* or deletions of 6p21.3 encompassing the *SYNGAP1* gene. Patients present with developmental delay, intellectual disability and ASD as well as epilepsy and sleep disturbances (Holder et al. 2019). PMD and *SYNGAP1*-ID are particularly relevant examples of rare monogenic neurodevelopmental disorders because, like idiopathic neurodevelopmental disorders, the phenotypic presentations are broad. Moreover, both are critical for development of excitatory neurotransmission, and imbalance between excitatory and inhibitory neurotransmission is one of the key findings in multiple neurodevelopmental disorders (Gao and Penzes 2015). Thus, it is important to understand the range and frequency of each of the symptoms associated with these diagnoses. We focus here on sensory features in these two disorders.

There are many tools available to assess sensory features, however the most common methods are caregiver surveys. The Sensory profile, developed in 1994, is a 99-item questionnaire of which 67 items were found to be uncommon amongst typically developing children (Dunn 1994). From the sensory profile, the Short-Sensory Profile (SSP) was developed which characterized behaviors in seven sensory areas (McIntosh et al. 1999). This was later updated to the Short Sensory Profile 2 (SSP-2, Dunn et al. 2014) based on the Dunn model of sensory processing (Dunn 1997). The SSP-2 evaluates overall scores in the sections of sensory and behavior. It also evaluates the four quadrants: seeking, avoiding, sensitivity and registration.

The most expansive exome sequencing study to date in individuals with Autism Spectrum Disorders has found that loss-of-function variants in *SHANK3* and *SYNGAP1* are among the most common monogenic etiologies for ASD (Satterstrom et al. 2020). Moreover, in clinical reports of cohorts of individuals with PMD and *SYNGAP1*-ID, both ASD and sensory features are commonly reported (Betancur and Buxbaum 2013; Jimenez-Gomez et al. 2019; Mieses et al. 2016). In a small study comparing SSP scores in 24 children with PMD to 61 children with non-syndromic ASD, PMD patients were found to have fewer sensory sensitivities but to score higher in the area of low energy (Mieses et al. 2016). Similarly, data from a patient registry found that 45 of the 48 patients with *SYNGAP1*-ID reported abnormal tactile responses, although these included both hypo-sensitivity manifested by decreased response to pain and hyper-sensitivity manifested by avoiding touch (Michaelson et al. 2018). However, the specific patterns of sensory processing differences in children with Phelan-McDermid Syndrome (PMD) and *SYNGAP1*-related intellectual disability (*SYNGAP1*-ID) remain unknown. Understanding these patterns is important for providing appropriate care to children with these disorders and for understanding the underlying mechanisms which cause their symptoms. Therefore, in this study we used the Short Sensory Profile 2 (SSP-2; Dunn 2014) to characterize sensory processing in patients with PMD and *SYNGAP1*-ID.

## Methods

Participants for this study was recruited from the Bluebird Clinic for Pediatric Neurology, Texas Children’s Hospital, and from the patient advocacy organizations Phelan McDermid Syndrome, Bridge the Gap: SYNGAP Education and Research Foundation, and SynGAP Research Fund, Inc. We provided the organizations with a recruitment letter that was sent out by email to their registry. This study was reviewed and approved by the Baylor College of Medicine Institutional Review Board for human studies.

### Participants

Adult caregivers of individuals age three and above with Phelan-McDermid Syndrome (PMD) or *SYNGAP1*-related intellectual disability (*SYNGAP1*-ID) were eligible to participate. Short sensory profile 2 responses were obtained from 41 subjects with PMD and 24 subjects with *SYNGAP1*-ID from December 2019 to June 2020.

### Procedures

This study used the Short Sensory Profile 2 (SSP-2; Dunn 2014). The SSP-2 is a 34-question parent survey in which respondents report the frequency of various sensory related behaviors. Questions are assigned to one of four sensory areas and scores from each area are summed to give a raw score in each of the four quadrants: seeking, avoiding, sensitivity and registration. Raw scores were compared to previously published standardized samples in which scores up to 1 standard deviation (SD) in either direction from the mean are considered just like the majority of others. Scores 1 SD to 2 SD are considered more than others and scores greater than 2 SD are much more than others. SSP-2 data was collected either in person or over the phone by MCM or JLH using the standard SSP-2 form (Dunn 2014). The survey was performed once per participant and was completed in 15-20 minutes. Participants answered almost always, frequently, half the time, occasionally, or almost never to each of the 34 questions which were then scored 1-5 (1 almost never, 5 almost always). Data were then transferred into an excel spreadsheet stored in a secure cloud location.

### Data Analysis

Raw total scores were calculated for each of the quadrants: seeking, avoiding, sensitivity and registration as well as the overall sensory and behavior scores. All scores were compared to previously published normative data (Dunn 2014). Histograms were used to display raw score distributions. Student’s unpaired t-test calculations were done in Excel. Correlations between each of the four quadrants were measured using R^2^ calculations in Matlab.

## Results

### PMD and SYNGAP1-ID patients perform outside the typical range on the Short Sensory Profile 2

We collected short sensory profile 2 (SSP-2) data from 41 individuals with PMD and 24 individuals with *SYNGAP1*-ID. The two groups were not significantly different except for age. The mean age for PMD patients was 12.9 years which was slightly higher than 8.08 years for *SYNGAP1*-ID (student’s t-test p = 0.02) (Table 1). Patient groups were similar in composition including sex, ethnicity and percent living in the United States.

We first looked at total sensory (Figure 1a) and behavior (Figure 1b) scores for patients with PMD and *SYNGAP1*-ID. These scores represent overall performance in the two main categories evaluated by the SSP-2. The majority of patients (37/41, 90% PMD; 21/24, 87.5% *SYNGAP1*-ID) scored above the range of typically developing responders (green shading) for sensory scores. All *SYNGAP1*-ID patients and all but one PMD patient scored above the expected scoring range (green shading) for behavior scores. There was no significant difference between PMD and *SYNGAP1*-ID patients in either scoring area (student’s unpaired t-test, Behavioral p = 0.08, Sensory p = 0.505). Thus, both PMD and *SYNGAP1*-ID patients exhibit more sensory and behavior differences than typically developing children but are not significantly different from each other.

**Figure 1.**
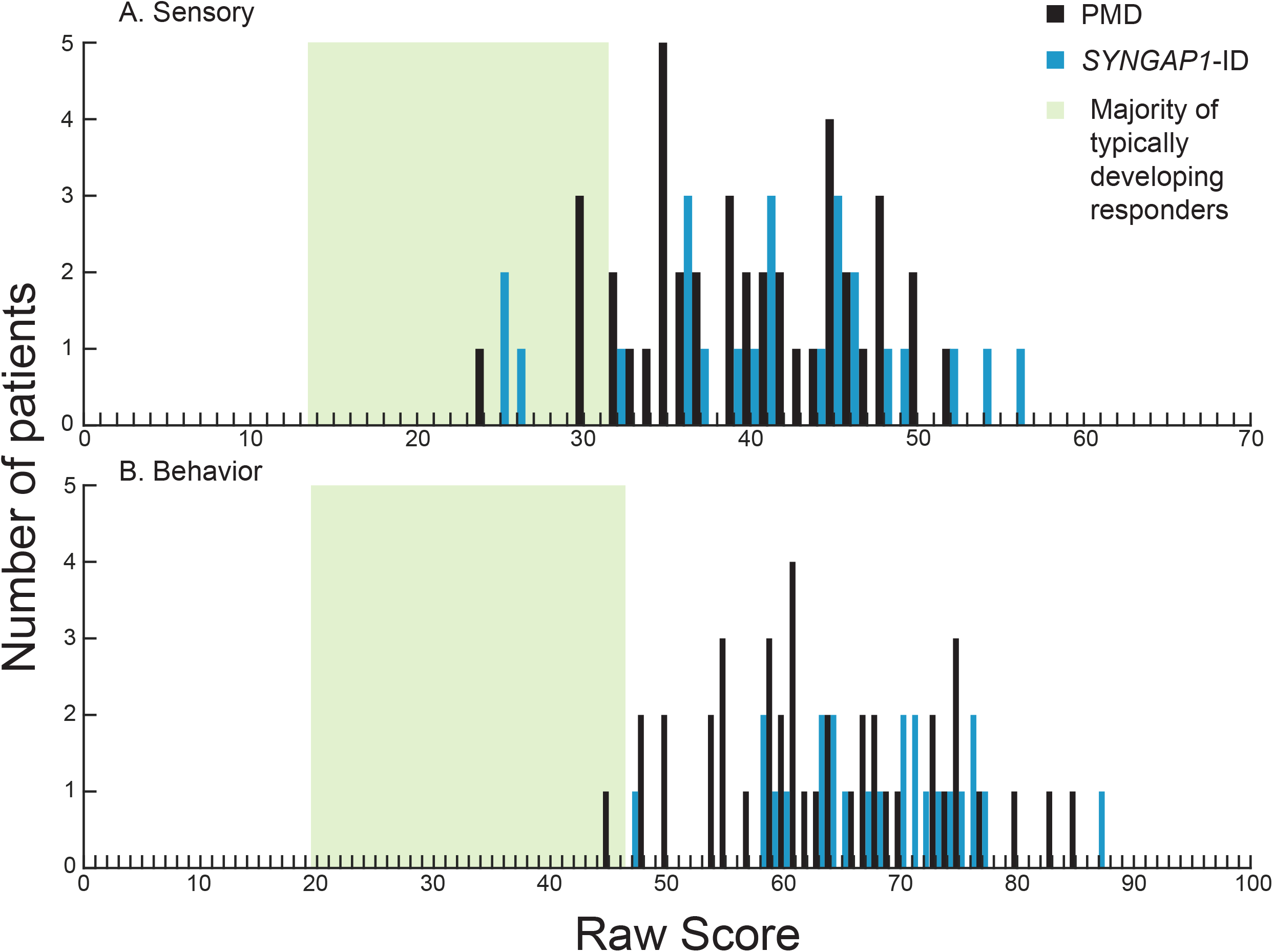
Distribution of scores in each of the two sections on the SSP-2 for PMD and *SYNGAP1*-ID Histograms showing number of PMD patients (black) and *SYNGAP1*-ID patients (blue) with each raw score in the sections of sensory (A) and behavioral (B). Green box indicates range of scores associated with the majority of typically developing responders based on prior validation of the SSP-2.

We next looked at the four quadrants measured by the SSP-2: seeking, avoiding, sensitivity and registration. These sub-categories provide more granular detail in the domains contributing to sensory behaviors. The majority of participants scored higher than most typically developing individuals (green shading, Dunn 2014) in all four areas (Figure 2). Specifically, in the areas of seeking, avoiding and registration, PMD and *SYNGAP1*-ID patients had overlapping score distributions that ranged from the same as most individuals to much more than most individuals (Figure 2a, b, d). Interestingly, in the sensitivity quadrant, scores from all PMD and *SYNGAP1*-ID patients in this study were more (9/41, 22% PMD; 5/24, 21% *SYNGAP1-*ID) or much more (32/41, 78% PMD; 19/24, 79% *SYNGAP1-*ID) than typically developing individuals (Figure 2c). When comparing the mean score for PMD patients to *SYNGAP1*-ID patients, *SYNGAP1*-ID patients scored significantly higher in the avoiding quadrant (29.46 vs 25.29, p = 0.004). There were no significant differences in mean score between PMD and *SYNGAP1*-ID patients in the other three areas (student’s unpaired t-test, seeking p = 0.37, sensitivity p = 0.91, registration p = 0.99). Thus, similarly to the overall scores, patients with PMD and *SYNGAP1*-ID exhibit more sensory features than typically developing children particularly in the area of sensory sensitivity. The notable difference in sensitivity is even more pronounced in the *SYNGAP1*-ID population.

**Figure 2.**
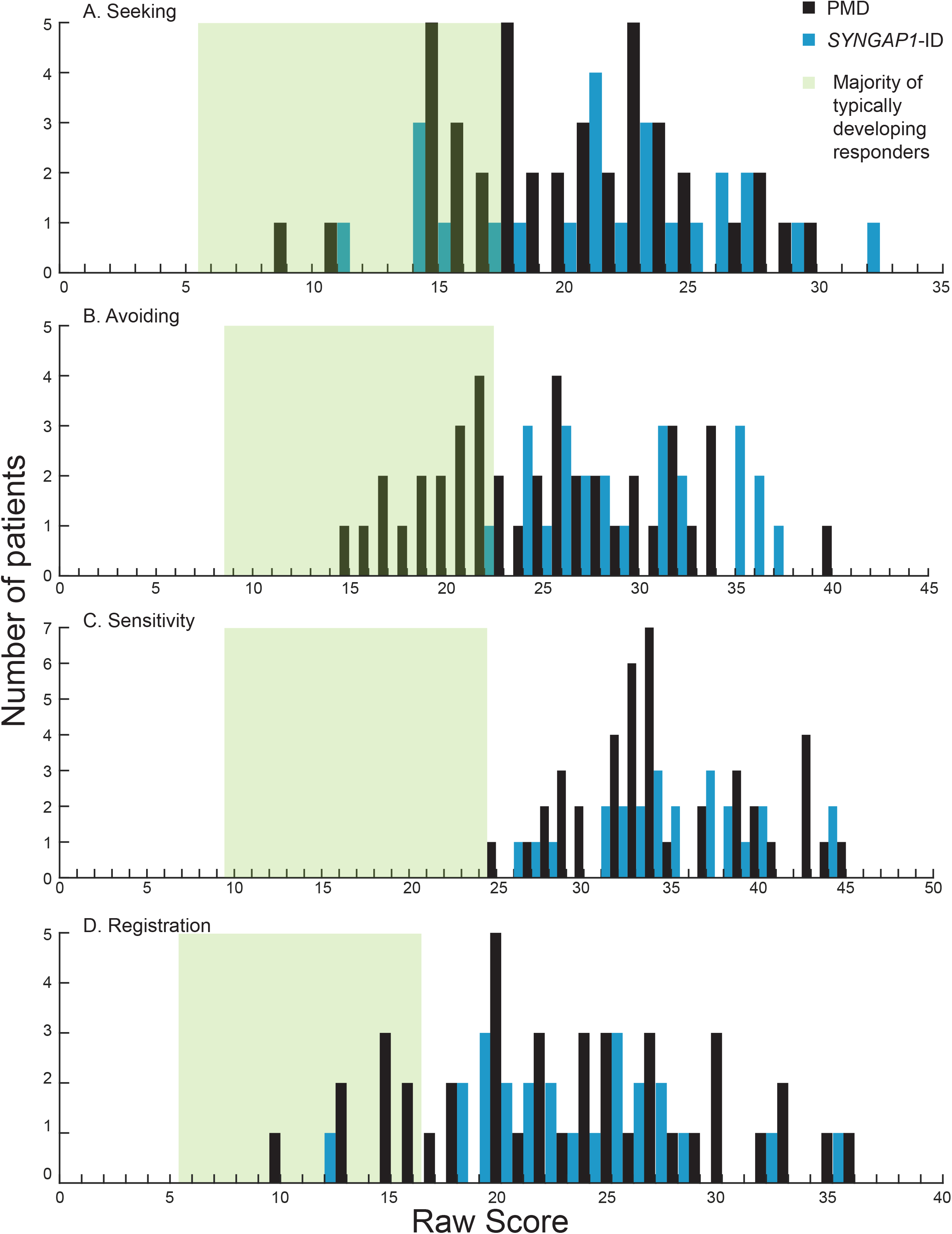
Distribution of scores in each of the four quadrants on the SSP-2 for PMD and *SYNGAP1*-ID Histograms showing number of PMD patients (black) and *SYNGAP1*-ID patients (blue) with each raw score in the quadrants of seeking (A), avoiding (B), sensitivity (C) and registration (D). Green box indicates range of scores associated with the majority of typically developing responders based on prior validation of the SSP-2.

### Both PMD and SYNGAP1-ID patients score high in avoiding and seeking

We noted that the majority of subjects scored in the “more than” or “much more than” range in all areas which was interesting because we had hypothesized that children who were higher in seeking would be lower in avoiding and vice versa. We therefore looked specifically at the distribution of scores in these two categories.

For PMD patients, we found that the most common combination was to be more than others in seeking and just like others in avoiding (7/41) (Figure 3a, red circle). 43% (18) patients were more or much more than others in both sensory patterns (Figure 3a, boxed area). Unlike for *SYNGAP1*-ID, there were five patients (12%) who scored just like typically developing children in both avoiding and seeking. There were also seven patients (17%) who exhibited more avoiding but were within the expected range for seeking and 11 patients (27%) who exhibited more seeking but were within the expected range for avoiding. (Figure 3a)

**Figure 3.**
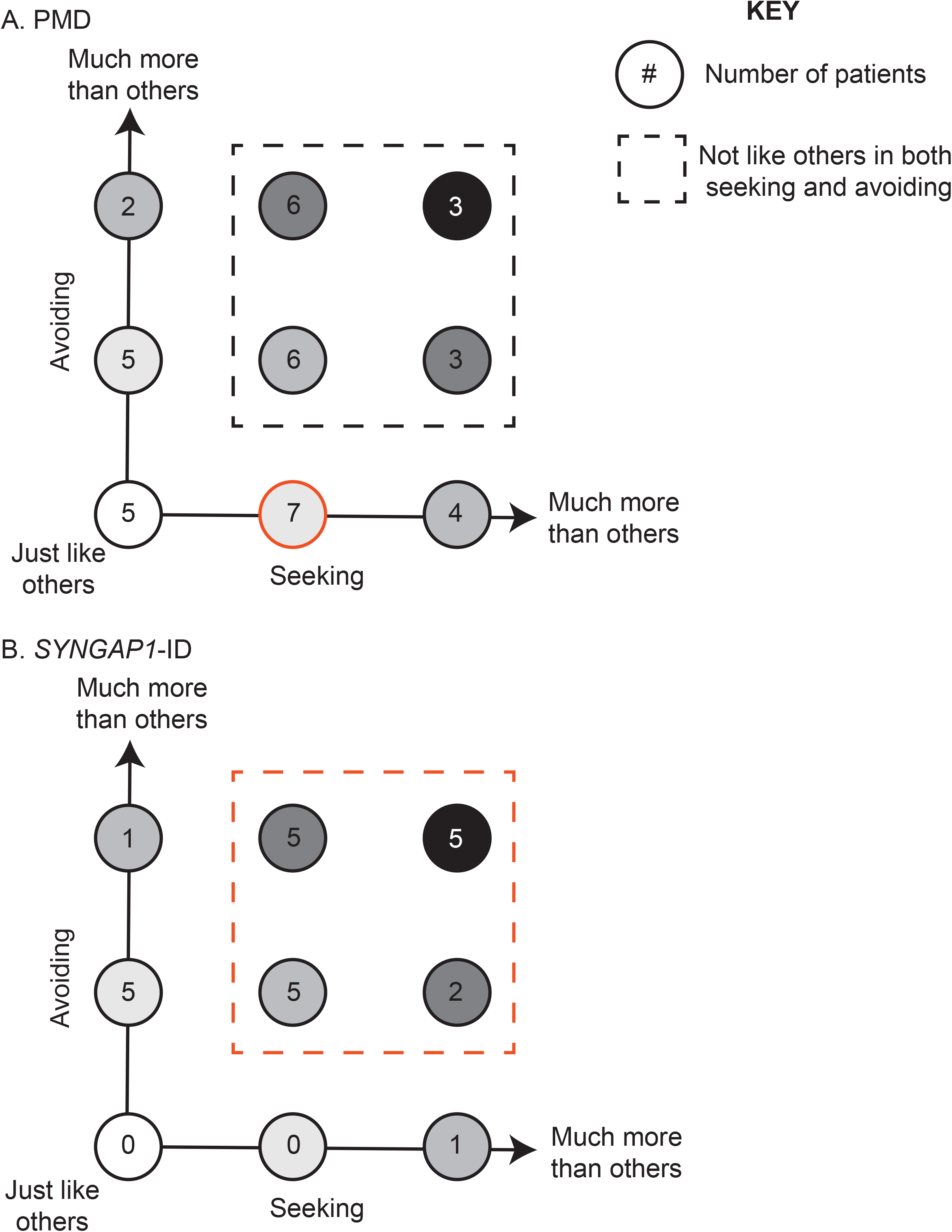
Association between seeking and avoiding scores Number of PMD (A) and *SYNGAP1*-ID (B) patients with each possible combination of seeking (x-axis) and avoiding (y-axis) scores. Each circle indicates a possible combination and is color coded based on severity. For example, white circle indicates patients scoring in the expected range for both areas with darker shading indicating higher scores. Subjects scoring outside the expected range in both categories indicated by dashed box. Most common combinations highlighted in red.

In contrast, for *SYNGAP1*-ID patients (Figure 3b), we found that the majority (71%) scored more or much more than others in both categories (Figure 3b, red dotted line). Specifically, five patients (21%) scored much more than others in both categories and five patients (21%) scored more than others in both categories. No patients scored in the just like others range for both features. Therefore, particularly in *SYNGAP1*-ID, patients exhibit a large number of atypical behaviors in both categories of seeking and avoiding.

### Registration and sensitivity scores are correlated in SYNGAP1-ID

We next asked if there was a correlation between scores in any of the four categories. Interestingly, *SYNGAP1*-ID patients demonstrated a moderate correlation between registration and sensitivity (R^2^ = .3703) that was also weakly present for PMD patients (R^2^ = .1565) (Figure 4a). A similar weak correlation was seen between avoiding and seeking scores in *SYNGAP1*-ID patients (R^2^ = .1822). In contrast, there was no correlation between scores for avoiding and seeking in PMD patients (R^2^ = .0517) (Figure 4b). Therefore, the specific patterns of sensory and behavioral features measured by the SSP-2 vary across patients. Having a high score in one area does not necessarily indicate a patient will have a high score in another area.

**Figure 4.**
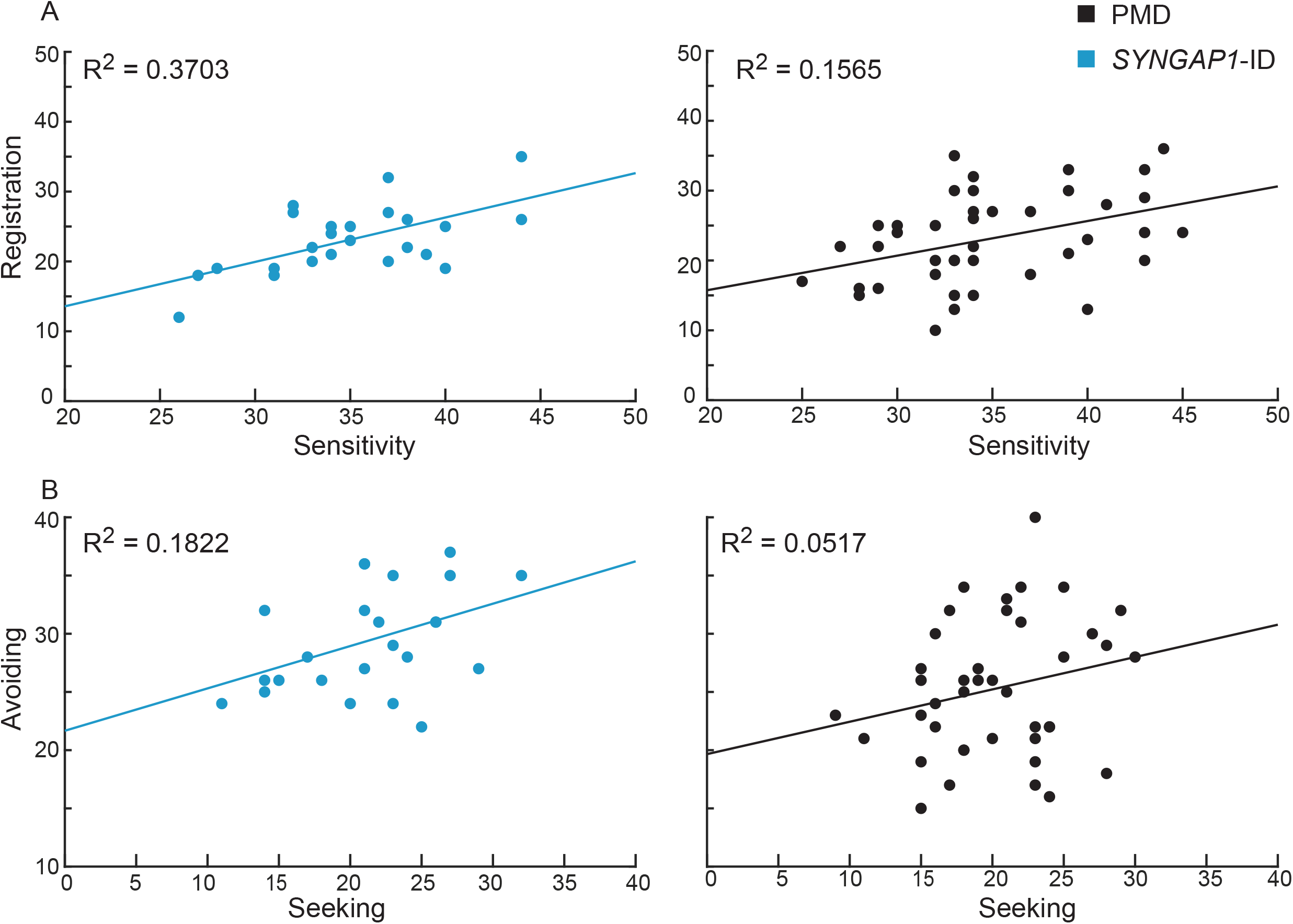
Correlation between sensory areas. Registration and sensitivity scores (A) for *SYNGAP1*-ID (left, blue) and PMD (right, black) were more strongly correlated than avoiding and seeking scores (B).

## Discussion

### Sensory Processing in PMD, SYNGAP1-ID and non-syndromic ASD

Significant work has demonstrated the prevalence of sensory differences in children with ASD using a variety of sensory survey tools (Leekam et al. 2007; Liss et al. 2006; Simpson et al. 2019). Recently, SSP-2 scores in ASD patients were reported to have a higher distribution in all four quadrants with 37-66% scoring in the “much more than others” range for each of the domains (Simpson et al. 2019). Interestingly, while Simpson et al reported ASD patients with scores ranging from much less than others through much more than others, we saw only expected range or higher scores in PMD and *SYNGAP1*-ID. This discrepancy could be related to our smaller patient sample size or could reflect the greater heterogeneity in non-syndromic ASD. Interestingly, when compared to children with Autism, sensory scores on the Short Sensory Profile (SSP) for patients with Fragile X syndrome exhibited similar ranges even when the mean score was significantly different (Rogers et al. 2003). Unlike the SSP-2, the SSP characterized children in seven areas: tactile, taste/smell, movement, and visual/auditory sensitivity as well as under-responsiveness/seeking, auditory filtering, and low-energy/weak. Thus, it is not possible to directly compare with our results. However, the narrower range of scores in our study may reflect differences between common and rare neurodevelopmental disorders.

Our study adds significantly to our limited prior understanding of sensory features in PMD and *SYNGAP1*-ID. Interestingly, scores for patients with PMD overlapped with those from patients with *SYNGAP1*-ID except in the area of avoiding. Children with *SYNGAP1*-ID scored higher in the avoiding quadrant indicating more frequent responses in this area. We also saw that patients from both groups scored highly in both seeking and avoiding. Finally, we found a correlation between sensitivity and registration in *SYNGAP1*-ID patients.

Sensory reactivity in PMD has been previously reported based on the SSP (Mieses et al. 2016). Interestingly, mean raw scores for PMD patients in Mieses et al were within the typical performance range for taste/smell and visual/auditory sensitivity. In contrast to our findings, Mieses et al found children with PMD to show more low energy/weak symptoms and *less* sensitivity when compared to children with idiopathic ASD. However, the SSP cannot be directly compared to the SSP-2. Therefore, further sensory evaluations in larger cohorts will be needed. Mouse models of PMD also demonstrate sensory deficits using tests of odor detection and odor discrimination (Drapeau et al. 2018). Olfaction is a common sensory feature to assay in mice because it is behaviorally relevant for these animals, however olfaction is rarely assayed or documented in the clinical setting (Sarnat and Flores-Sarnat 2020). Our study is the first prospective evaluation of sensory profiles in patients with *SYNGAP1*-ID. A recent *SYNGAP1*-ID international conference that included both affected individuals with caregivers and providers concluded that sensory processing is affected in nearly 100% of patients (Weldon et al. 2019). Similarly, review of a *SYNGAP1*-ID registry database identified 48 entries with information on sensory processing. Of these, 45 patients exhibited features of sensory impairment with a particular emphasis on abnormal tactile responses in 20 entries (Michaelson et al 2018). These findings are consistent with the high sensory scores observed for *SYNGAP1*-ID patients in our study. Intriguingly, multiple investigations have discovered that mice with mutations in *Syngap1* display impaired sensory processing including sensory-motor gating (Guo et al. 2009), hypersensitivity to capsaicin-induced thermal hypernociception (Duarte et al. 2011) and tactile discrimination (Michaelson et al. 2018). Additionally, mouse studies suggest sensory processing deficits are due to decreases in cortical synaptic connectivity (Michaelson et al. 2018). If sensory processing differences are related to impairments in sensory-motor gating or nociception, this could explain why *SYNGAP1*-ID patients were selectively higher in the area of avoiding. Future studies should focus on mechanisms of sensory processing in the mouse model as this maybe a valuable target for testing novel targeted therapeutics.

### How can patients have high scores in both seeking and avoiding

We were surprised to find that both PMD and *SYNGAP1*-ID patients scored higher than expected in the areas of both seeking and avoiding. These high scores were not due to patients being high in one or the other, but in fact most patients were high in both categories. In reviewing the literature, we find that several previous studies have identified similar types of clusters in non-syndromic ASD. For example, Lane et al identified 5 clusters using the short sensory profile including a cluster defined by high under-responsivity and sensory seeking (Lane et al. 2011). Similarly, Liss et al used selected questions from the sensory profile with additional sensory specific questions and found 4 clusters including a cluster with high scores in under-responsivity and sensory seeking (Liss et al. 2006).

We also noted a correlation between sensitivity and registration in *SYNGAP1*-ID patients. Specific association between sensitivity and avoiding have been reported for non-syndromic ASD (Simpson et al. 2019) but we are not aware of previous correlations between registration and sensitivity using the SSP-2. As the SSP-2 is used to characterize cohorts from other types of syndromic ASD, it will be interesting to see if this correlation is common or unique.

### Implications for future research

Characterizing sensory profiles of children with PMD and *SYNGAP1*-ID is important for providing appropriate care and for understanding the underlying mechanisms of the disorders. Further, quantification of sensory metrics can serve as an end point for evaluating the efficacy of treatments in clinical trials. Given the significant deviation in scores in all four areas from the majority of typically developing children seen in our cohort, we propose that the SSP-2 can be used to quantitatively measure improvement in children with PMD or *SYNGAP1*-ID during future clinical trials.

## Data Availability

Raw data is available upon reasonable request

## Acknowledgements

The authors would like to thank and acknowledge the patients and families who contributed to this study.

## Declarations

### Ethics Approval

The questionnaire and methodology for this study was approved by the Human Research Ethics committee of Baylor College of Medicine (Ethics approval number: H-45748).

### Consent to Participate

Written informed consent was obtained from the parents. All information was de-identified and reported as averages.

## Figure Caption Sheet

*Table 1*. 41 participants with PMD and 24 participants with *SYNGAP1*-ID were included in this study.

## Notes

### Competing Interest Statement

The authors have declared no competing interest.

### Funding Statement

Funded in part by Bridge the Gap: SYNGAP1 Education and Research Foundation

### Author Declarations

Baylor College of Medicine Institutional Review Board

