## Supplementary material for "Brief Report: Sensory processing phenotypes in Phelan-McDermid Syndrome and *SYNGAP1*-related Intellectual Disability": Table I

**Table 1: Demographics**

|  | **PMD** | ***SYNGAP1*-ID** |
| --- | --- | --- |
| N | 41 | 24 |
| Male | 18 (44%) | 12 (50%) |
| Age in years | 12.9 (3-46) | 8.08 (0-19) |
| Ethnicity |  |  |
| Caucasian | 36 (88%) | 20 (83%) |
| Hispanic | 3 (7%) | 3 (12.5%) |
| American Indian | 1 (2.5%) | 0 (0%) |
| African American | 0 (0%) | 1 (4%) |
| Unknown | 1 (2.5%) | 0 (0%) |
| Living in the US | 31 (76%) | 21 (87.5%) |
